# Trends in Cognitive Function Pre- and Post- Stroke: Finding from the China Health and Retirement Longitudinal Study

**DOI:** 10.1101/2021.09.24.21263136

**Authors:** Jianian Hua, Jianye Dong, Yueping Shen

## Abstract

**Introduction:** The magnitude of changes in cognitive function after stroke among the Chinese population is not clear. We aimed to learn the trajectories of cognitive function before and after incident stroke among Chinese participants.

**Methods:** Data were obtained from a nationally representative study. A total of 13311 Chinese participants aged 45 years or older and without a history of stroke were assessed at baseline between June 2011 and March 2012 and in at least one cognitive test between Wave 2 to Wave 4. Cognitive function was assessed by a global cognition score, which included episodic memory, visuospatial abilities, and executive function. A linear mixed model was developed to explore the repeated measurements.

**Results:** During the seven-year follow-up, we observed 610 (4.6%) participants experienced the first stroke. The baseline mean (SD) age was 58.6 (9.2) years. 47.3% of them were male. There was no difference in the baseline cognitive function and decline rate of pre-stroke cognition between stroke survivors and stroke-free participants after adjusting for covariates. Among the stroke survivors, the acute decline was -0.123, -0.169, and -0.135 SD/y in cognitive domains of episodic memory, visuospatial ability, and global cognition, respectively. The executive function did not decline acutely after stroke. In the years following stroke, the decline rate of executive function was 0.056 SD/y faster than the rate before stroke. The decline rate of episodic memory, visuospatial ability, and global cognition did not accelerate after stroke.

**Conclusion:** Before stroke onset, the cognitive function of Chinese stroke survivors was similar to that of stroke-free participants. Incident stroke was associated with acute decline in global cognition, episodic memory, and visuospatial abilities, and accelerated decline in orientation, attention, and calculation abilities. The cognitive trajectories revealed by our study highlights the need to care for the cognitive decline among Chinese stroke survivors.

## 1 Introduction

Dementia or cognitive impairment is common after stroke^1-3^. A meta-analysis conducted in 2021 reported that around 10% to 30% of patients had dementia after stroke^4^. In developed countries, population ageing coupled with a decline in mortality after stroke is bringing with an even higher prevalence of post-stroke cognitive impairment (PSCI)^5-7^. Elucidating the cognitive trends pre- and post-stroke would be helpful to determine impact of acute stroke treatment on cognitive function and to design health policy to manage the burden of PSCI^8^. However, since most studies were hospital-based, the trends of cognitive function before stroke remained unclear. Baseline cognition, an important risk factor, is impossible to measure. Methods such as Informant Questionnaire of Cognitive Decline in the Elderly (IQCODE) or medical records from health system could achieve information on pre-stroke cognitive function^9,10^. Nevertheless, they have two important limitations. The first is recall bias^11^. Second, these data were converted into dichotomous variables. This prevented researchers from observing the linear trends before and after stroke.

Several articles had reported the cognitive trend pre- and post-stroke. A 2015 *JAMA* article included over twenty thousand England participants, finding that stroke survivors experienced a faster rate of cognitive decline after stroke than before stroke^12^. A 2021 *STOKE* article examined American participants from three study epochs, and they further reported the cognitive score of stroke survivors declined faster even before the stroke, compared with that of the stroke-free participants^13^. To the best of our knowledge, the cognitive trends mentioned above has not been investigated among Chinese populations. Furthermore, there is still room for improvement of methods used to calculate the trend.

In 2021, the China Health and Retirement Longitudinal Study (CHARLS), a nationally representative database, published its fourth survey and offered us the opportunity to explore the cognitive trends across multiple surveys. Therefore, the aim of our study was to investigate the temporal pattern of trends in cognitive function before and after stroke among Chinese populations.

## 2 Methods

### 2.1 Study population

CHARLS is a community-based and nationally representative cohort study in China and a sister study of the Health and Retirement Study (HRS). The baseline (Wave 1) was conducted 2011 and included 17708 randomly selected Chinese participants from 150 country units distributed in 28 provinces of China via multistage probability sampling^14^. Data from Wave 1 (2011), Wave 2 (2013), Wave 3 (2015), and Wave 4 (2018) was used in the current analysis.

Figure 1 showed the flow chart of sample selection. 16040 individuals completed cognitive tests at baseline. 352 participants less than 45 at baseline were excluded. We excluded 381 participants with brain damage or mental retardation and 170 individuals with memory-related disease at baseline. Of the 15137 remaining participants, 423 with a history of stroke at baseline were excluded. 42 individuals missed covariates at baseline. We retained participants who received at least one cognitive test during Wave 2 to 4, excluding 1361 participants lost to follow-up. Finally, 610 stroke survivors and 12701 participants without incident stroke were included^15^.

**Figure 1.**
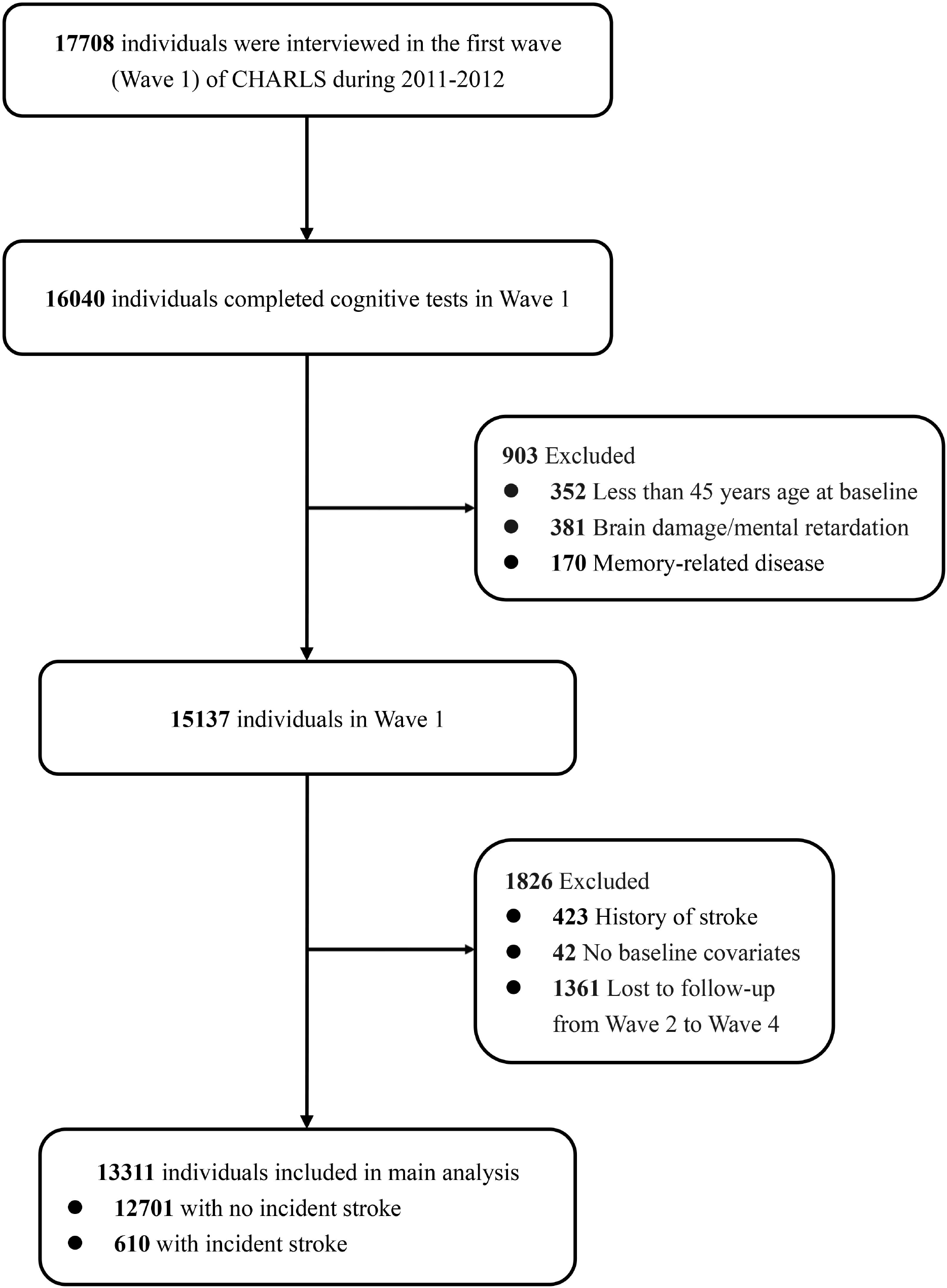
Flow chart of participant selection.

### 2.2 Assessment of cognitive function and stroke

Cognitive assessments were conducted in all four waves and included three domains. Its reliability and validity have been demonstrated in previous studies^16-18^. Immediate and delayed (5 minutes) recall tests of ten Chinese nouns were used to assess episodic memory. 0.5 points for each correct word. The score of episodic memory ranged from 0 to 10. Visuospatial ability was assessed by asking participants to redraw a figure. One score was received for success. The executive function was assessed by asking participants to repeatedly subtract 7 from 100 and to identify the date, season, and the day of the week. It assessed participants’ ability of calculation, attention, and orientation. The score for executive function could range from 0 to 10^19^.

We created z-scores for each test using the mean and SD of scores at baseline. The global cognition z scores were created by standardizing the sum of the three individual z-scores in the same manner^20^.

We evaluated stroke as a modifier of the association between neighborhood and mortality. The first stroke was based on a time-updated self- or proxy-report of a doctor’s diagnosis (“Has a doctor ever told you that you had a stroke?”). No information on transient ischemic attacks, stroke subtypes, or stroke severity was available. For some stroke participants, the diagnosis time was recorded by interview. For others, the time was defined as the midpoint between the date of the last wave without stroke history and that of the wave reporting a stroke history.

### 2.3 Covariates

Covariates related to stroke and cognitive function were selected, including age, sex, education, marital status, residential area, smoking, drinking, hypertension, dyslipidemia, diabetes, cancer, lung diseases, heart problems, depression, and the number of instrumental activities of daily living IADLS^15^. Education was classified as illiterate, primary school, middle school, and high school, and above. Marital status was classified married and the other status. The 10-item Center for Epidemiologic Studies Short Depression Scale (CES-D-10) was used to measure depressive symptoms. This score can range from 0 to 30. A score ≥12 was defined as depression^21^.

### 2.4 Statistical analysis

Baseline characteristics and baseline cognitive scores were compared between participants who did and did not experience a stroke during the follow-up, and between those included in the analysis and those who lost to follow-up.

Linear mixed models were used to fit the trends of cognitive function. It accounts for between-group variation and within-group correlation of repeated outcomes^22,23^. In model A, random effects were estimated for intercept and time since baseline. Fixed effects were estimated for intercept, time (years since baseline), stroke (yes or no), time*stroke interaction, stroke status (yes or no), and covariates. Intercept and time variables showed the baseline cognition and cognitive change rate of those without stroke, respectively. The coefficient of the “stroke” variable accounts for the baseline difference between those who did and did not experience a stroke during the follow-up. The stroke*time interaction evaluated the difference cognitive change rates of participants with and without stroke. The “stroke status” was a time-varying dichotomies variable, changed from “0” to “1” at the time of stroke onset, reflecting the acute cognitive decline after stroke. Random effects were estimated for intercept and time. Model B included variates in Model A and added stroke*stroke status*time-after-stroke interaction. Time after stroke were fitted for both fixed effects and random effects. This interaction indicated changes in cognitive slope before and after stroke. Then, the stroke*time interaction reflected the difference between the rate of change from baseline to incident stroke and that from baseline to end of follow-up^24^.

We calculated participant-specific predicted scores for a 70-year-old female with the most common baseline characteristics (Figure 2). Random effects were set to zero.

**Figure 2.**
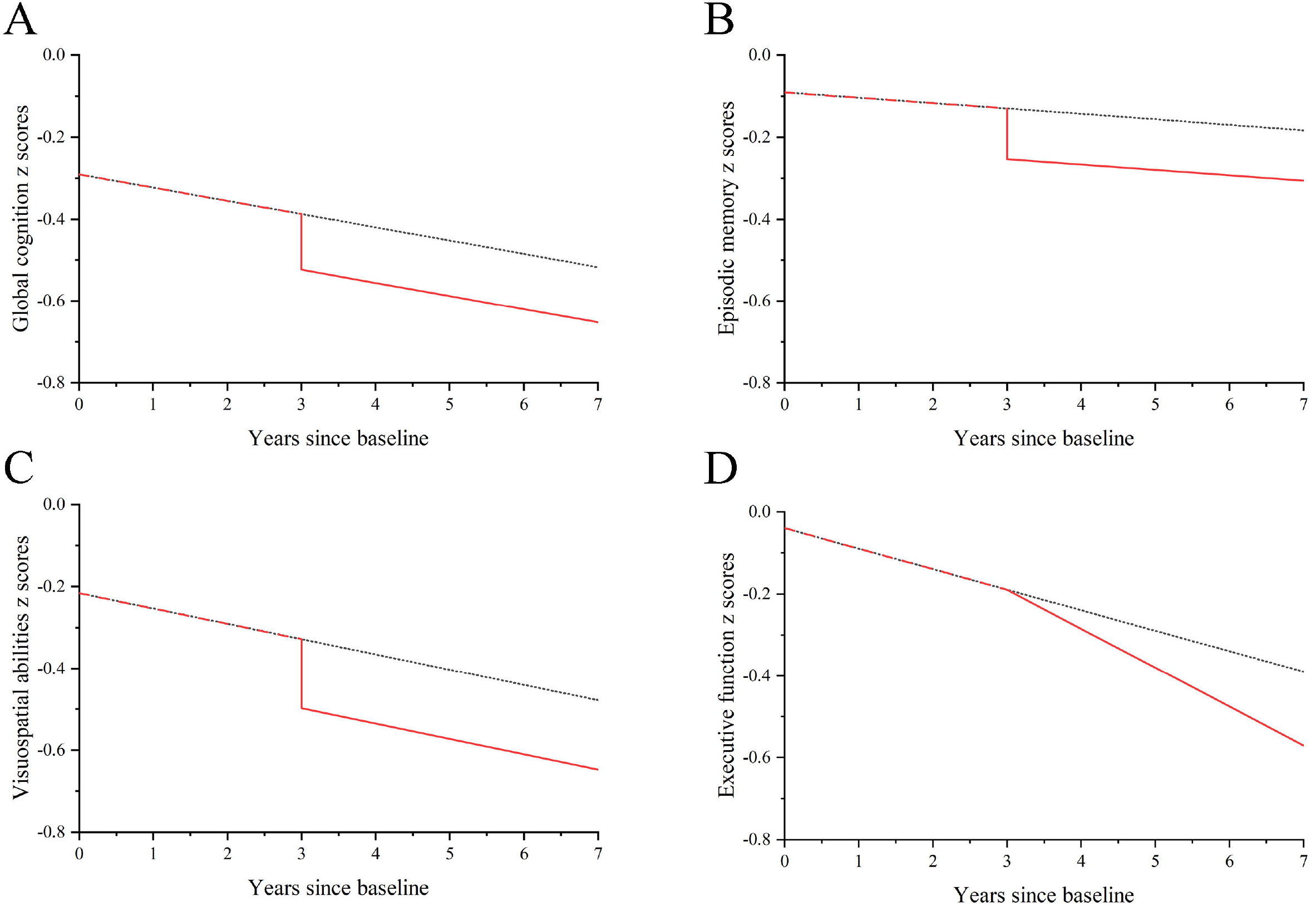
Predicted changes in cognitive z scores among all participants during the follow-up. The black dotted curves represented participants without incident stroke. The red dashed curves represented the pre-stroke cognitive trajectories among participants with incident stroke. The red sloid curves represented the post-stroke trajectories. The predicted values of cognitive score were calculated for a 70-year-old female. Her education level was primary school. She lived in rural area, and was married, but without current smoking, current drinking, hypertension, dislipidiemia, diabetes, cancer, lung diseases, heart problems, or depression. Her IADL score was zero. She experienced one incident stroke at the end of the third year.

P values were 2-sided, with 0.05 being the threshold for statistical significance. All analyses were performed using SAS version 9.4 (SAS Institute Inc., Cary, NC, USA).

### 2.5 Sensitivity analysis

In sensitivity analysis, we restricted our subjects to participants (both with and without stroke) who received tests in all four waves, this more closely showed the trends in relatively healthier. We included participants who experienced a stroke before baseline^12^.

## 3 Results

Among the 13311 participants who were included in the main analysis, 610 (4.6%) experienced at least one incident stroke during the seven-year follow-up. The baseline characteristics was shown in Table 1. The mean ± age of individuals without incidence stroke was 58.6 ± 9.2 years. 47.3% of them were male. 66.4% of them hadn’t finished middle school. 78.8% lived in rural areas.

**Table 1.** Baseline characteristics of participants with or without incident stroke during follow-up.

|  | No incident stroke<br>(n = 12701) | Incident stroke<br>(n = 610) | P Value<sup>a</sup> | Adjusted P value<sup>b</sup> |
| --- | --- | --- | --- | --- |
| Continuous variables, mean (SD) |  |  |  |  |
| Age | 58.6 (9.2) | 61.0 (8.8) | <0.001 | - |
| Number of IADLs | 0.2 (0.6) | 0.3 (0.8) | <0.001 | <0.001 |
| Global cognition scores | 10.6 (4.3) | 10.0 (4.1) | <0.001 | 0.114 |
| Episodic memory scores | 3.3 (1.9) | 3.1 (1.8) | <0.001 | 0.452 |
| Visuospatial ability scores | 0.6 (0.5) | 0.6 (0.5) | 0.0033 | 0.054 |
| Executive function scores | 6.6 (2.8) | 6.3 (2.9) | <0.001 | 0.109 |
| Categorical variables, n (%) |  |  |  |  |
| Males | 5925 (47.3) | 290 (48.3) | 0.335 | - |
| Education |  |  | <0.001 | 0.247 |
| Illiterate | 3293 (26.3) | 173 (28.8) |  |  |
| Primary school | 5026 (40.1) | 258 (42.9) |  |  |
| Middle school | 2665 (21.3) | 104 (17.3) |  |  |
| High school and above | 1556 (12.4) | 66 (11.0) |  |  |
| Marital status |  |  | <0.001 | <0.001 |
| Married | 11156 (89.0) | 520 (86.5) |  |  |
| Other status | 1384 (11.0) | 81 (13.5) |  |  |
| Residential area |  |  | 0.285 | 0.002 |
| Urban | 2660 (21.2) | 122 (20.3) |  |  |
| Rural | 9880 (78.8) | 479 (79.7) |  |  |
| Current smoking | 4861 (38.8) | 254 (42.3) | <0.001 | <0.001 |
| Current drinking | 3201 (25.2) | 165 (27.5) | 0.034 | <0.001 |
| Hypertension | 2731 (21.8) | 245 (40.8) | <0.001 | <0.001 |
| Dyslipidemia | 1041 (8.30) | 89 (14.8) | <0.001 | <0.001 |
| Diabetes | 633 (5.1) | 51 (8.5) | <0.001 | <0.001 |
| Cancer | 111 (0.9) | 3 (0.5) | 0.0462 | 0.0790 |
| Lung Diseases | 1228 (9.8) | 68 (11.3) | 0.015 | <0.001 |
| Heart problems | 1384 (11.4) | 107 (17.8) | <0.001 | <0.001 |
| Depression | 2579 (20.6) | 165 (27.5) | <0.001 | <0.001 |

<sup>a</sup>Calculated using ANOVA for continuous covariates and $\chi^2$ test for categorical covariates.
<sup>b</sup>Calculated using generalized linear models for continuous covariates and logistic regression for categorical covariates, after adjustment for baseline age and sex.

Compared with participants without incident stroke, those with incident stroke were older; had a high number of IADLs; had higher percentages of unmarried, smoking, drinking, hypertension, dyslipidemia, diabetes, lung disease, heart problems, and depression. There was no difference in sex proportions between the two groups. After adjusting for age and sex, the stroke group tended to live in rural areas. The stroke group had lower cognitive scores in all domains. After adjustment for age and sex, there was no significant difference in baseline cognitive function between participants with and without incident stroke.

Following our hypothesis mentioned above, the model reflected these trajectories of cognitive function: (1) the predicted baseline score of the without-stroke group (Intercept), (2) the decline rate of the without-stroke group (Slope), (3) the difference in baseline score (Difference in intercept), (4) the difference in decline rate in the stroke group compared to the without-stroke group in the pre-stroke period (Difference in slope before stroke), (5) the acute cognitive change after stroke among the stroke group, and (6) how much the decline rate change after stroke among the stroke group.

As shown in Table 2 and Figure 2, there was no difference in the baseline cognitive scores between the two groups. For individuals who remained stroke-free, their cognitive scores consistently declined in all cognitive domains. Likewise, the cognitive function of participants with stroke declined before stroke onset. The decline rates were similar across the two groups. Therefore, we hadn’t observed pre-stroke cognitive disadvantage of people with stroke compared to people without stroke.

**Table 2.** Trajectories of cognitive function among all participants from 2011 and 2018^a^.

|  | Global cognition |  |  |  | Episodic memory |  |  |  | Visuospatial ability |  |  |  | Executive function |  |  |  |
| --- | --- | --- | --- | --- | --- | --- | --- | --- | --- | --- | --- | --- | --- | --- | --- | --- |
|  | Model A |  | Model B |  | Model A |  | Model B |  | Model A |  | Model B |  | Model A |  | Model B |  |
|  | β<br>(95% CI) | P | β<br>(95% CI) | P | β<br>(95% CI) | P | β<br>(95% CI) | P | β<br>(95% CI) | P | β<br>(95% CI) | P | β<br>(95% CI) | P | β<br>(95% CI) | P |
| Variables |  |  |  |  |  |  |  |  |  |  |  |  |  |  |  |  |
| Baseline age | -0.018<br>(-0.020, -0.017) | <0.001 | -0.018<br>(-0.020, -0.017) | <0.001 | -0.021<br>(-0.023, -0.020) | <0.001 | -0.021<br>(-0.023, -0.020) | <0.001 | -0.012<br>(-0.013, -0.011) | <0.001 | -0.012<br>(-0.013, -0.011) | <0.001 | -0.009<br>(-0.010, -0.007) | <0.001 | -0.009<br>(-0.010, -0.007) | <0.001 |
| Intercept of<br>without-stroke<br>group | 0.221<br>(0.114, 0.328) | <0.001 | 0.221<br>(0.115, 0.328) | <0.001 | 0.540<br>(0.435, 0.646) | <0.001 | 0.541<br>(0.435, 0.647) | <0.001 | 0.213<br>(0.107, 0.318) | <0.001 | 0.212<br>(0.107, 0.318) | <0.001 | 0.174<br>(0.072, 0.277) | <0.001 | 0.175<br>(0.073, 0.278) | <0.001 |
| Difference in<br>intercept<sup>b</sup> | 0.017<br>(-0.048, 0.082) | 0.606 | 0.017<br>(-0.050, 0.085) | 0.613 | 0.059<br>(-0.012, 0.130) | 0.106 | 0.045<br>(-0.030, 0.119) | 0.241 | -0.002<br>(-0.077, 0.074) | 0.969 | 0.014<br>(-0.065, 0.093) | 0.736 | 0.012<br>(-0.050, 0.074) | 0.700 | -0.021<br>(-0.087, 0.044) | 0.520 |
| Slope of without-stroke group | -0.032<br>(-0.035, -0.030) | <0.001 | -0.032<br>(-0.035, -0.030) | <0.001 | -0.013<br>(-0.016, -0.010) | <0.001 | -0.013<br>(-0.016, -0.010) | <0.001 | -0.037<br>(-0.041, -0.034) | <0.001 | -0.037<br>(-0.041, -0.034) | <0.001 | -0.050<br>(-0.052, -0.048) | <0.001 | -0.050<br>(-0.052, -0.048) | <0.001 |
| Difference in slope<br>before stroke<sup>c</sup> | -0.005<br>(-0.023, 0.011) | 0.507 | -0.006<br>(-0.026, 0.014) | 0.556 | -0.010<br>(-0.030, 0.009) | 0.295 | -0.004<br>(-0.026, 0.018) | 0.731 | -0.001<br>(-0.023, 0.022) | 0.948 | -0.008<br>(-0.032, 0.017) | 0.546 | -0.008<br>(-0.025, 0.008) | 0.308 | 0.008<br>(-0.011, 0.027) | 0.422 |
| Acute change after<br>stroke<sup>d</sup> | -0.138<br>(-0.227, -0.049) | 0.002 | -0.135<br>(-0.228, -0.042) | 0.004 | -0.139<br>(-0.239, -0.039) | 0.006 | -0.123<br>(-0.227, -0.019) | 0.021 | -0.143<br>(-0.258, -0.029) | 0.014 | -0.169<br>(-0.292, -0.047) | 0.007 | -0.086<br>(-0.176, 0.004) | 0.060 | -0.056<br>(-0.148, 0.035) | 0.230 |
| Changes in slope<br>after stroke | None | None | -0.003<br>(-0.035, 0.029) | 0.857 | None | None | -0.019<br>(-0.052, 0.014) | 0.257 | None | None | 0.022<br>(-0.017, 0.062) | 0.271 | None | None | -0.045<br>(-0.073, -0.017) | 0.001 |
| Log likelihood | -47397.5 |  | - 47396.35 |  | - 57312.65 |  | - 57310.1 |  | - 58379.45 |  | - 58377.15 |  | - 53,235.95 |  | - 53,227.95 |  |

\*All coefficients and confidence intervals were shown in the form of z score.
<sup>a</sup>Adjusted for baseline age (shown in Line “Baseline age”), sex, education, marital status, residential area, current smoking, current drinking, hypertension, dyslipidemia, diabetes, cancer, lung diseases, heart problems, depression, and number of IADLs.
<sup>b</sup>The difference in intercept in the stroke group compared to the without-stroke group.
<sup>c</sup>The difference in slope rate in the stroke group during pre-stroke period compared to the without-stroke group during the whole follow-up period.
<sup>d</sup>The amount the cognitive scores changed at the “stroke point” among the stroke group, measured as the first post-stroke value minus the last pre-stroke value.

**Table 3.** Changes in slope after incident stroke compared with slope before incident stroke^a,b^.

| | $\beta$ (95% CI) | P value |
| --- | --- | --- |
| Global cognition | -0.074 (-0.093, -0.054)* | <0.001 |
| Episodic memory | -0.043 (-0.063, -0.023) | 0.034 |
| Visuospatial ability | 0.027 (0.005, 0.049) | 0.215 |
| Executive function | -0.061 (-0.078, -0.045) | <0.001 |

\*All coefficients and confidence intervals were shown in the form of z score.
<sup>a</sup>Adjusted for age, sex, education, marital status, residential area, current smoking, current drinking, hypertension, dyslipidemia, diabetes, cancer, lung diseases, heart problems, depression, and number of IADLs.
<sup>b</sup>The coefficient reflected the amount of the slope changed after incident stroke among the stroke group.

The acute cognitive decline after score was shown in Table 2 and the vertical line in Figure 2. Stroke survivors showed a significantly acute decline in episodic memory (β, -0.123 SD; 95% CI, -0.176, -0.70; P=0.021; Model B), visuospatial ability (β, -0.169 SD; 95% CI, -0.232, -0.106; P=0.007; Model B), and global cognition (β, -0.135 SD; 95% CI, -0.183, -0.135; P=0.004; Model B). We did not detect acute cognitive decline after incident stroke in executive function (β, -0.056 SD; 95% CI, -0.103, -0.010; P=0.230; Model B).

In the years following stroke, all cognitive domains declined over time (Figure 3). Executive function declined 0.045 SD/y faster than it did before stroke (β, -0.045 SD; 95% CI, -0.059, -0.031; P=0.001; Model B). No accelerated decline rate was observed in episodic memory, visuospatial ability, or global cognition, compared with the pre-stroke rate.

We conducted sensitivity analysis after excluding participants who didn’t receive cognitive tests in all four waves and included participants with baseline stroke, respectively. The results remained similar.

## 4 Discussion

In this national cohort of Chinese participants aged :245 years, we observed an accelerated cognitive decline in global cognition after stroke. There was no significant difference in baseline cognition and pre-stroke cognitive decline rate between participants with and without stroke. For episodic memory abilities, visuospatial abilities, and global cognition, stroke patients experienced an acute cognitive decline at the time of stroke and showed the same decline rate compared with their pre-stroke rate. For calculation, orientation, and attention abilities, stroke participants didn’t show an immediate cognitive decrement at stroke onset but experienced a steeper cognitive decline in the long term after stroke.

Our results were partly in agreement with the findings from other ethnicities. To the best of our knowledge, only participants from two countries were examined the cognitive trajectories before and after stroke. Reasons for Geographic and Racial Differences in Stroke (REGARDS)^12,25^ and English Longitudinal Study of Ageing (ELSA)^15^, namely, were from the UK. Health and Retirement Study (HRS)^13,26,27^ and Chicago Health and Aging Project (CHAP)^26^, namely, were from America. The cognitive tests in REGARDS assessed the following function: recall and temporal orientation, new learning, verbal memory, and executive function. A study from the REGARDS reported acute cognitive decline and persistent cognitive decline in recall and temporal orientation, and verbal memory abilities. New learning abilities declined acutely after stroke but remained a same change rate after stroke compared with the pre-stroke rate. Consistent with our results, the executive function of their participants didn’t decline immediately at the time of stroke, but experienced accelerated cognitive decline but no persistent cognitive decline in hadn’t examined the difference in the pre-stroke cognitive decline between participants with incident stroke and the natural cognitive decline among control group^12^. The ELSA tested three cognitive domains, including memory, semantic fluency, and orientation. For UK participants in ELSA, all cognitive domains declined acutely at stroke onset and declined faster in the long periods after stroke. HRS only tested memory abilities and reported similar findings. The Americans in HRS experienced an acute cognitive decrement immediately after stroke. The decline rate after stroke was not significantly faster than that before stroke^13,27^. CHAP reported an accelerated cognitive decline in the years after stroke. However, the authors didn’t consider the effect of acute cognitive decline, which would lead to bias^26^. Usage of z-score makes results across studies comparable. The acute cognitive decline in memory among the Chinese population (−0.123 SD) was similar to that among UK population (−0.150 SD), and less than that among the American population in Epoch 3 of HRS (−0.25 SD)^13,15^.

A key difference between our participants and the English in ELSA or the Americans in HRS lied on the baseline cognitive scores and the pre-stroke cognitive trend. Studies using ELSA and HRS found stroke survivors had lower baseline cognition and faster cognitive decline before stroke, compared with the stroke-free participants. This is different from our findings in all cognitive domains. The study learning HRS analyzed stroke survivors, stroke decedents, and stroke-free group, separately. Not surprisingly, the pre-stroke cognitive decline rate was much faster among stroke decedents^13^. This might be due to that stroke patients have higher comorbidity of cardiovascular risk factors, such as hypertension, diabetes, and atherosclerosis, during quite a significant time before stroke onset. These risk factors could accelerate cognitive decline even after they were adjusted in the multiple regression^28^. Most of our stroke participants were “stroke survivors” (only 12 of them died during the follow-up). Nevertheless, stroke survivors in HRS also had lower pre-stroke cognitive scores. The study learning ELSA hadn’t taken the effect of fatal stroke into account^15^. We don’t know whether the results would change after considering the subgroup.

The acute cognitive decline could be explained by mechanisms of cognitive impairment in the acute and subacute stage of stroke, including infarction of the brain tissue, hypoperfusion, and “misery perfusion”, which was represented by decreased cerebral blow flow and increased oxygen extraction fraction in areas distal to the lesions^11,29,30^. Mechanisms explaining the long-term cognitive decline after stroke remained to be elucidated. Incident stroke could trigger or accelerate the Alzheimer-disease-related pathologies, such as amyloid deposition^31^. And, neurodegeneration ahead of stroke could exacerbate the brain injury after stroke^32,33^. Stroke survivors had a higher prevalence of hypertension, diabetes, and atrial fibrillations. The vascular-related comorbidities and immune response contributed to the cognitive deficits after stroke^28,34^. Cerebral small vessel disease, reflected by whiter matter hyperintensity, lacune, perivascular space, and microbleeds, was associated with greater cognitive decline in general cognition^35,36^. Recurrent stroke, clinical or subclinical, might increase the risk of dementia in the long term after stroke^3,32^. However, we didn’t have the image data at the stroke onset and follow-up waves.

The most advantage of this article was our study population. We firstly reported how the cognitive trajectories differed before and after incident stroke among Chinese participants. Previous studies learning only stroke survivors missed pre-stroke cognitive tests and might not generalize to the Chinese population. In recent years, the mortality of stroke in China is decreasing due to improved medical technology and treatment^37,38^, for example, the growing use of tissue-type plasminogen activator(r-tPA) and standard therapy. Now China have over 300 Comprehensive Stroke Centers and over 1000 Tertiary Stroke Centers. In recent decades, a growing number of observational studies and reports have reported the prevalence and types of cognitive or emotional disfunctions after stroke in developing countries^39^. In this regard, studies in China are dedicated to not only motor disfunction, but also complications such as cognitive decline. The cognitive trajectories revealed by our study highlights the needs to care the acute cognitive decline in cognitive function among Chinese stroke survivors. Compared with the previous studies, we also improved the usage of linear mixed models. Our models considered the trajectories of stroke-free group and stroke group simultaneously.

However, our study had several limitations. The first limitation was the lack of medical records, leading to a serious of deficits. We were unable to differentiate stroke subtypes, including cerebral infarction, intracerebral hemorrhage, and transient ischemic attacks. Since ischemic stroke accounts for approximately 80% of stroke, our results more likely reflected conditions in ischemic stroke survivors. Furthermore, the self-reported or the investigator-reported stroke status would introduce recall bias. An American study showed self- and proxy-reported stroke history had about a sensitivity of 74% and a specificity of 93% to detect stroke^40^. The situation in our cohort was unknown. To compare the pre- and post-stroke cognitive trend, specificity, which was much higher, was more important than sensitivity. Some of our participants lack data on recurrent stroke, which prevented us to conduct sensitivity analysis. Insufficient data on stroke severity, location, or acute treatment prevented us from controlling for more factors^9^. Second, our cohort didn’t record the cause of death. Hence, we were unable to identify stroke decedents from those who suffered a stroke and soon died in one wave. Participants who experienced a severe stroke or was aphasia after stroke were more likely to die or drop out, and they tended to have poorer health status at baseline. This might reduce our ability to detect the full effects of stroke on cognitive trends, especially trends before stroke^41^. Third, although we controlled for quite a number of confounders, we cannot rule out the possibility of residual confounders, such as atrial fibrillation, body mass index (BMI), and Apolipoprotein E (APOE) genotype^42^.

## 5 Conclusion

Our study added to the understanding of the cognitive trajectories after stroke among Chinese stroke patients. It highlighted the need for improved cognitive assessment in stroke trials. Future studies could learn the mechanisms for the acute and accelerated long-term cognitive decline, and explore modifiable risk factors and effective interventions to reduce the cognitive burden after stroke.

## Data Availability

The data used in this manuscript were obtained from the CHARLS. Prof. Yaohui Zhao (National School of Development of Peking University), John Strauss (University of Southern California), and Gonghuan Yang (Chinese Center for Disease Control and Prevention) are the principal investigators.

## 9 Author Contributions

Jianian Hua contributed to the conception and design of the study. Jianian Hua and Yueping Shen organized the database. Jianian Hua and Jianye Dong performed the statistical analysis. Jianian Hua and Jianye Dong wrote the first draft of the manuscript.Yueping Shen reviewed the manuscript. All authors approved the final version of the paper.

## 11 Acknowledgements

We thank the China Center for Economic Research and the National School of Development of Peking University for providing the data.

## 12 Data Availability Statement

The data used in this manuscript from the China Health and retirement Longitudinal Study (CHARLS). We applied the permission for the data access (http://charls.pku.edu.cn/zh-CN) and got the access to use it. Prof. Yaohui Zhao (National School of Development of Peking University), John Strauss (University of Southern California), and Gonghuan Yang (Chinese Center for Disease Control and Prevention) are the principal investigators.

